# Risk of SARS-CoV-2 reinfection 18 months after primary infection: population-level observational study

**DOI:** 10.1101/2022.02.19.22271221

**Authors:** Maria Elena Flacco, Graziella Soldato, Cecilia Acuti Martellucci, Giuseppe Di Martino, Roberto Carota, Antonio Caponetti, Lamberto Manzoli

**Affiliations:** Department of Environmental and Prevention Sciences, University of Ferrara, 44121 Ferrara, Italy; Local Health Unit of Pescara, 65124 Pescara, Italy; Department of Medical and Surgical Sciences, University of Bologna, 40100 Bologna, Italy

## Abstract

Current data suggest that SARS-CoV-2 reinfections are rare, but uncertainties remain on the duration of the natural immunity, its protection against Omicron variant, finally the impact of vaccination to reduce reinfection rates. In this retrospective cohort analysis of the entire population of an Italian Region, we followed 1,293,941 subjects from the beginning of the pandemic to the current scenario of Omicron predominance (up to mid-January 2022). After an average of 334 days, we recorded 260 reinfections among 84,907 previously infected subjects (overall rate: 0.31%), two hospitalizations (2.4 x100,000), and one death. Importantly, the incidence of reinfection did not vary substantially over time: after 18-22 months from the primary infection, the reinfection rate was still 0.32%, suggesting that protection conferred by natural immunity may last beyond 12 months. The risk of reinfection was significantly higher among the unvaccinated subjects, and during the Omicron wave.

## Introduction

A number of field studies reported low rates of SARS-CoV-2 reinfections after a primary episode ^1,2^. Uncertainties remain, however, on the duration of natural immunity, its protection against BA.1/B.1.1.529 (Omicron) variant, and the impact of vaccination to decrease reinfection rates. We performed a retrospective cohort study on the entire population of an Italian region in order to estimate the incidence of SARS-CoV-2 reinfections and diseases according to vaccination status, predominant viral strain, and time after primary infection.

## Methods

We included all residents in the Abruzzo Region, Italy with ≥1 positive nasopharyngeal swab detected by the regional accredited laboratories, from pandemic start (March 2, 2020) up to November 30, 2021. On January 14, 2022 (to allow ≥45 days of follow-up), we extracted all data of the official vaccination, COVID-19, demographic, hospital and co-pay exemption datasets of the National Healthcare System, merging individual information through encrypted fiscal code ^3^.

A reinfection was defined by the presence of two positive RT-PCR samples detected ≥45 days apart with ≥1 intermediate negative RT-PCR test ^4,5^. Subjects were classified as “vaccinated” if they received ≥1 dose of BNT162b2, ChAdOx1 nCoV-19, mRNA-1273 or JNJ-78436735, ≥14 days before the reinfection. The analyses were stratified by demographic and clinical characteristics, time after primary infection (0-5, 6-11, 12-17, 18-22 months), and predominant circulating variant (pre-Omicron vs. Omicron; from Dec. 28, 2021 on). A two-sided *P*-value<.05 was considered significant. Stata, version 13.1 (Stata Corp.) was used for all analyses.

## Results

From the start of the pandemic, a total of 152,986 infections were detected among the 1,293,941 residents in the Abruzzo Region. After the exclusions of the subjects with a follow-up <45 days, or lacking negative intermediate swabs, a total of 84,907 subjects with a primary infection were included in the analysis. The average time after the primary infection was 334 days; 2829 subjects had a follow-up longer than 18 months (Table 1).

**Table 1.**
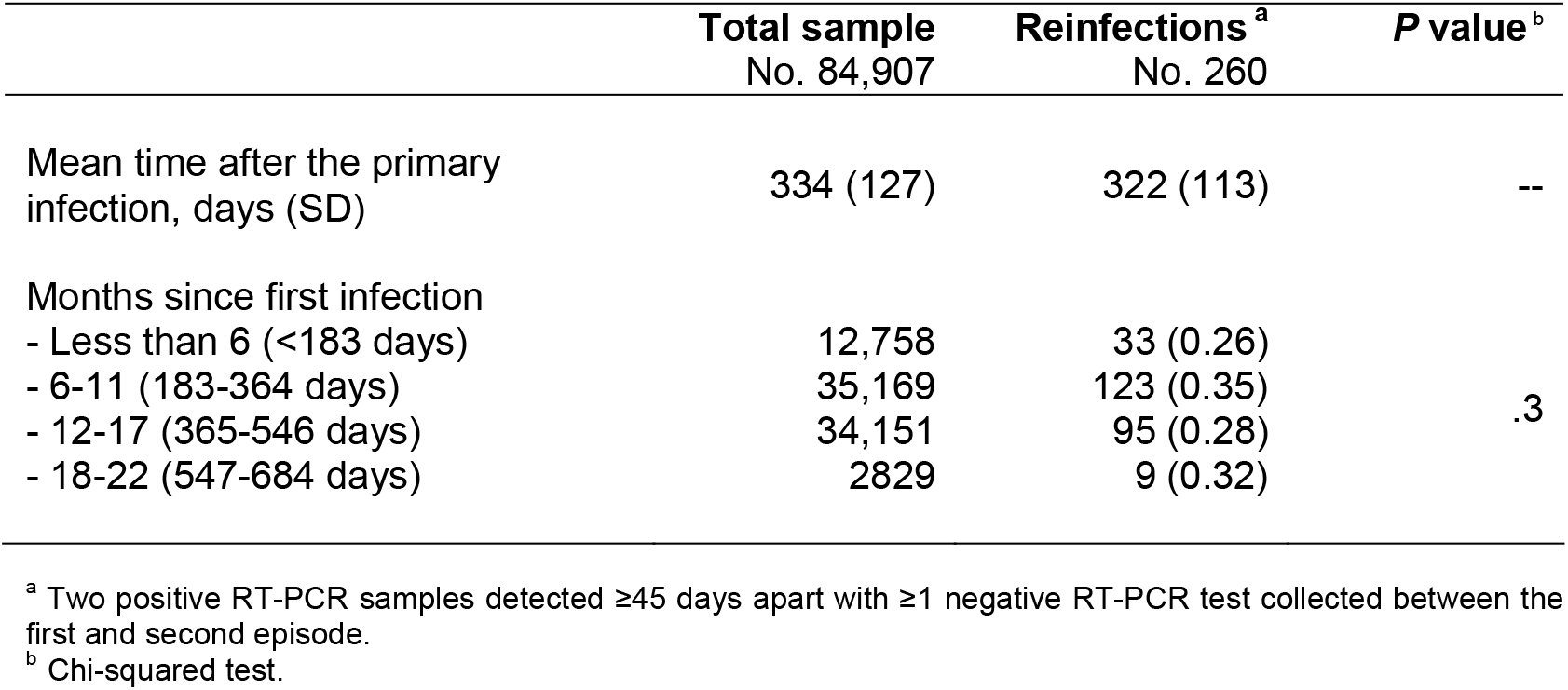
Time after the primary SARS-CoV-2 infection.

Overall, the incidence of reinfection was 0.31% (n=260). Two subjects were hospitalized due to COVID-19 (2.4×100,000), and one died (a 73-year old female, with major cardiovascular disease and diabetes).

As shown in Table 2, the reinfection rate was significantly higher among females, younger subjects, and unvaccinated individuals (0.50% vs. 0.25% among those who received ≥2 vaccine doses). Moreover, a markedly higher rate of reinfections was recorded during the first 17 days of the Omicron wave (n=144/84,791; 8.5 per day) than during the 317 days of the pre-Omicron period (n=116/84,907; 0.4 per day). In contrast, the incidence of reinfection did not vary substantially by baseline comorbidities and over time: after 18 or more months from the primary infection (up to 22 months), the reinfection rate was still 0.32%. The multivariable analysis (Cox proportional hazards model adjusted for all the recorded variables, included a priori) confirmed univariate results (Table 2). When the analysis was stratified by circulating variant, the effectiveness of ≥2 vaccine doses was substantially higher in pre-Omicron (adjusted hazard ratio – HR vs. unvaccinated: 0.20; 95% Confidence Interval: 0.12-0.33) than Omicron wave (HR: 0.57; 0.36-0.90).

**Table 2.** SARS-CoV-2 reinfections, overall and by selected covariates.

| Variables | Total sample No. | Reinfections<sup>a</sup> No. (%) | P value<sup>b</sup> | Reinfection Adj. HR (95% CI) |
| --- | --- | --- | --- | --- |
| Overall | 84,907 | 260 (0.31) | -- | -- |
| Age class, years |  |  |  |  |
| 60+ | 19,846 | 25 (0.13) |  | 1 (ref. cat.) |
| 30-59 | 38,801 | 114 (0.29) | <.001 | 0.72 (0.55-0.95) |
| 0-29 | 26,260 | 121 (0.46) |  | 0.30 (0.18-0.50) |
| Mean age in years (SD) | 43.1 (22.3) | 34.3 (19.3) | <.001 | 0.99 (0.98-0.99) |
| Gender |  |  |  |  |
| - Females | 43,099 | 160 (0.37) |  | 1 (ref. cat.) |
| - Males | 41,808 | 100 (0.24) | <.001 | 0.62 (0.48-0.80) |
| Diabetes<sup>c</sup> |  |  |  |  |
| - No | 80,669 | 250 (0.31) |  | 1 (ref. cat.) |
| - Yes | 4238 | 10 (0.24) | .4 | 1.57 (0.79-3.12) |
| Hypertension<sup>c</sup> |  |  |  |  |
| - No | 75,730 | 245 (0.32) |  | 1 (ref. cat.) |
| - Yes | 9177 | 15 (0.16) | .009 | 0.96 (0.51-1.79) |
| Cardiovascular diseases<sup>c</sup> |  |  |  |  |
| - No | 76,294 | 246 (0.32) |  | 1 (ref. cat.) |
| - Yes | 8613 | 14 (0.16) | .011 | 0.88 (0.47-1.65) |
| COPD<sup>c</sup> |  |  |  |  |
| - No | 81,856 | 253 (0.31) |  | 1 (ref. cat.) |
| - Yes | 3051 | 7 (0.23) | .4 | 0.95 (0.44-2.04) |
| Kidney diseases<sup>c</sup> |  |  |  |  |
| - No | 83,607 | 260 (0.31) |  | 1 (ref. cat.) |
| - Yes | 1300 | 0 (0.0) | .044 | 0 (NE) |
| Cancer<sup>c</sup> |  |  |  |  |
| - No | 81,618 | 257 (0.31) |  | 1 (ref. cat.) |
| - Yes | 3289 | 3 (0.09) | .023 | 0.40 (0.13-1.27) |
| COVID-19 Vaccination<sup>d</sup> |  |  |  |  |
| - No | 21,926 | 110 (0.50) |  | 1 (ref. cat.) |
| - 1 dose | 35,995 | 82 (0.23) | <.001 | 0.43 (0.32-1.58) |
| - 2 or more doses | 26,986 | 68 (0.25) |  | 0.34 (0.25-0.47) |

Adj.: adjusted; HR: hazard ratio; CI: confidence interval; SD: standard deviation; ref. cat.: reference category; NE: not estimable.
<sup>a</sup> Two positive RT-PCR samples detected $\geq 45$ days apart with $\geq 1$ negative RT-PCR test collected between the first and second episode.
<sup>b</sup> Chi-squared test.
<sup>c</sup> Subjects with the selected comorbidities in the Regional co-pay exemption database (Italian "Esenzioni Ticket" file) or an hospital admission in the last ten years (from the Italian SDO database of administrative discharge abstracts) with the following ICD-9-CM codes in any diagnosis field—250.xx (diabetes); 401.xx-405.xx (hypertension); 410.xx-412.xx or 414.xx-415.xx or 428.xx or 433.xx-436.xx (major cardiovascular or cerebrovascular diseases - CVD); 491.xx-493.xx (chronic obstructive pulmonary disease - COPD); 580.xx-589.xx (kidney diseases); 140.xx-172.xx or 174.xx-208.xx (cancers).
<sup>d</sup> At least 14 days before the reinfection.

## Discussion

This study confirms and expands previous findings reporting a low risk of SARS-CoV-2 reinfection, and a very low risk of severe or lethal COVID-19 for those who recovered from a primary infection ^2,6^, suggesting that the protection conferred by the natural immunity lasts beyond 12 months. Although the marked increase of the reinfection rates during the Omicron wave is concerning, the risk of a secondary severe disease or death remained close to zero. Therefore, despite the vaccines were able to significantly reduce the likelihood of reinfection in both pre-Omicron and Omicron waves, the risk-benefit profile of multiple vaccine doses for this population should be carefully evaluated.

## Data Availability

All data produced in the present study are available upon reasonable request to the corresponding author.

## Authors’ contributions

Concept and design: M.E.F., G.S.; C.A.M., L.M.

Acquisition, analysis, or interpretation of data: G.S., G.D.M., R.C., L.M.

Drafting of the manuscript: M.E.F., C.A.M., L.M.

Critical revision of the manuscript for important intellectual content: G.S., G.D.M., A.C., L.M.

Statistical analysis: M.E.F., L.M.

Administrative, technical, or material support: G.S., G.D.M., R.C., A.C.

Supervision: A.C., L.M.

L.M. had full access to all the data in the study and takes responsibility for the integrity of the data and the accuracy of the data analysis. All data produced in the present study are available upon reasonable request to the corresponding author.

## Ethics Committee

The study was conducted according to the guidelines of the Declaration of Helsinki, and the research protocol was submitted to the Ethics Committee of the Emilia-Romagna Region, which gave ethical approval for this work (protocol code 2525/21).

